# Epigenomic prediction of cardiovascular disease risk and interactions with traditional risk metrics

**DOI:** 10.1101/19006965

**Authors:** Kenneth Westerman, Alba Fernández-Sanlés, Prasad Patil, Paola Sebastiani, Paul Jacques, John M. Starr, Ian Deary, Qing Liu, Simin Liu, Roberto Elosua, Dawn L. DeMeo, José M. Ordovás

## Abstract

Epigenome-wide association studies for cardiometabolic risk factors have discovered multiple loci associated with incident cardiovascular disease (CVD). However, few studies have sought to directly optimize a predictor of CVD risk. Furthermore, it is challenging to train multivariate models across multiple studies in the presence of study- or batch effects. Here, we analyzed existing DNA methylation data collected using the Illumina HumanMethylation450 microarray to create a predictor of CVD risk across three cohorts: Women’s Health Initiative, Framingham Heart Study Offspring Cohort, and Lothian Birth Cohorts. We trained Cox proportional hazards-based elastic net regressions for incident CVD separately in each cohort, and used a recently-introduced cross-study learning approach to integrate these individual predictions into an ensemble predictor. The methylation-based risk score (MRS) predicted CVD time-to-event in a held-out fraction of the Framingham dataset (HR per SD = 1.28, p = 2e-3) and predicted myocardial infarction status in the independent REGICOR dataset (OR per SD = 2.14, p = 9e-7). These associations remained after adjustment for traditional cardiovascular risk factors and were similar to those from elastic net models trained on a directly merged dataset. Additionally, we investigated interactions between the MRS and both genetic and biochemical CVD risk, showing preliminary evidence of an enhanced predictive power in those with less traditional risk factor elevation. This investigation provides proof-of-concept for a genome-wide, CVD-specific epigenomic risk score and suggests that the DNA methylation data may enable the discovery of high-risk individuals that would be missed by alternative risk metrics.

## Introduction

DNA methylation is an important epigenetic pathway through which genetic variants and environmental exposures impact disease risk^1,2^. Methylation at specific cytosine-phosphate-guanine (CpG) sites has been associated with disease in epigenome-wide association studies, even showing associations in blood as a convenient but non-target tissue such as for type 2 diabetes^3^. Methylation-based risk scores allow genome-wide aggregation of epigenetic information, similarly to the more established genetic risk scores, and allow for the use of models with arbitrary complexity. These risk scores are often developed initially by using methylation as a proxy for disease risk factors, such as body mass index (BMI)^4^ and general aging-related morbidity^5^. Alternatively, given sufficient sample size, epigenetic associations with disease risk can be modeled directly^6^.

Associations between DNA methylation and cardiovascular disease (CVD) have been explored in many different cohorts and using diverse approaches. Cross-sectional associations have been found across multiple relevant tissues, namely blood, aorta, and other vascular tissues^7^. Some investigations aimed at cardiovascular risk factors have discovered CpGs predictive of CVD development^8,9^, while Mendelian randomization approaches have suggested causality of at least some of these CpG-risk factor associations^10^. A few studies directly modeling incident CVD as a primary outcome have either been conducted using only global (not locus-specific) methylation levels^11^, or have found limited additional predictive power in the presence of known risk factors^12^. A recent large-scale meta-analysis found multiple CpG sites predictive of incident coronary heart disease, but focused on univariate approaches^13^. We have previously investigated methylation regions and modules associating with incident CVD, generating mechanistic insights but without aggregating these results into a direct predictor of risk^14^. Additionally, it is unclear how the CVD risk tracked by DNA methylation is redundant with or complementary to existing risk metrics, including genetic scores^15^ and those based on traditional cardiovascular risk factors (e.g. the Framingham Risk Score for generalized CVD)^16^.

Combining signal across population-scale cohorts can increase sample size while attenuating the effect of study-specific biases and confounding factors, but can be prone to emergent sources of confounding from “batch” effects or other systematic biases in methylation data across cohorts. This is especially problematic when there is notable class imbalance (i.e. different outcome frequencies) across cohorts^17^. The most common method for dealing with this heterogeneity is meta-analysis, but standard meta-analysis approaches are restricted to univariate (one CpG site at a time) models. Other approaches include batch effect correction on the input dataset (e.g. ComBat^18^), direct adjustment for batch/study in linear models, or adjustment for derived variables intended to capture technical biases (e.g. surrogate variable analysis^19^), but these approaches can often lead to over- or under-estimates of true biological effects^17^. An alternative approach described recently, cross-study learning, instead trains an ensemble predictor consisting of one or multiple models per cohort^20^. This strategy allows the use of arbitrarily complex models while avoiding technical confounding from direct combination of the datasets.

In order to develop an improved DNA methylation-based cardiovascular risk predictor using multiple training cohorts, we used a cross-study learning method to develop an ensemble of penalized time-to-event regression risk models. The resulting composite risk score performed well in a held-out data subset, predicting survival even in the presence of traditional risk factors, and showing similar performance to models trained on naively merged datasets. External validation was achieved in a case-control for prevalent myocardial infarction (MI). Further, interactions were assessed between the composite methylation-based risk score and other risk predictors, finding a potentially enhanced prediction in those with low Framingham Risk Scores.

## Methods

### Study participants and phenotype collection

Phenotypes (demographic, anthropometric, biochemical, and clinical), DNA methylation data, and imputed genotypes were available either from publicly available controlled-access databases or upon request from the cohorts. Cohort-specific details are provided in Supplementary methods. Blood-based biochemical markers (total cholesterol, LDL-cholesterol, HDL-cholesterol, triglycerides, fasting glucose, high-sensitivity C-reactive protein, and systolic blood pressure) were log10-transformed for all analyses. In the Lothian Birth Cohort 1936, LDL was estimated from total cholesterol and triglycerides using the Friedewald equation. Diabetes was defined as either use of diabetes medication or a measured fasting blood glucose level of >125 mg/dL. Median imputation was used to fill missing values for diabetes, medication use, and smoking status (thus assuming no diabetes, no medication use, and no smoking where these values were missing). Analysis of these datasets was approved by the Tufts University Health Sciences Institutional Review Board (protocol 12592), and all subjects gave informed consent.

### DNA methylation data processing

DNA methylation data for all initial cohorts (WHI, FHS, and LBC) were collected using the Illumina HumanMethylation450 microarray platform^21^ and downloaded as raw intensity files. Standard preprocessing steps were performed for each cohort, including sample-wise and probe-wise filters, background correction, and probe type normalization. After quality control and filtering steps, 390597 CpG sites were shared between the 3 datasets, formatted as beta values (ratio of methylated signal to total microarray signal). Preprocessing details are described in the Supplementary Methods.

### CVD risk prediction modeling

Study-specific CVD risk prediction models were trained using penalized Cox proportional hazards regressions with the elastic net penalty. CVD events were defined as above, and times were right-censored based on the most recent exam available in each cohort. The elastic alpha parameter was set at 0.05, based on prior observations of good performance on Illumina methylation microarray datasets^22^, and the penalty parameter *λ* was optimized through 5-fold cross-validation. For each model, only the most variable 100,000 CpGs according to median absolute deviation (∼25% of all available sites shared across platforms) were included in order to decrease the computational burden and ensure that the selected CpGs would have meaningful interindividual variation.

The cross-study learner (CSL) was constructed as an ensemble of study-specific regression models. Predictions from each single-study learner (SSL) were combined using the “stacking” approach^20^, implemented as follows. First, predictions from each SSL to both itself and the other training datasets were combined into a design matrix (with dimensions *N*_*total*_ x # SSLs). This formed the input to an additional penalized Cox regression (ridge regression with *λ* optimized through 5-fold CV and coefficients restricted to be non-negative) of all training studies at once. Coefficients from this regression, corresponding to input study-specific SSLs, were normalized to sum to one to produce the CSL weights. For prediction in new datasets, SSL predictions were each standardized to mean zero and unit variance before calculating their weighted sum (using the “stacking” weights) as the final CSL score.

A series of approaches for combining information across cohorts were tested as alternatives to the CSL. The naive “combined” approach consisted of simply aggregating observations from all training sets into a single dataset and training an elastic net regression as described above while adjusting for study as a fixed effect. The ComBat method trained across all studies as with the “combined” approach, but included an empirical Bayes-based preprocessing step to directly adjust the dataset for study differences while preserving variation along the “axis” of incident CVD events^18^.

MRS evaluation in FHS was performed using Cox proportional hazards models, with a series of models adjusting for covariates including demographics, anthropometrics, biochemical values, and cell subtype estimates. Robust standard errors were used to account for family structure as has been suggested for clustered data^23^ and used for epigenetic risk models in FHS^24^. The proportional hazards assumption was confirmed (p > 0.05) using the *cox*.*zph* R function. To compare risk scores generated using different models (combined and ComBat-preprocessed) to the CSL, Cox regressions adjusting for the “basic” covariate set were used to evaluate the CSL MRS alone, the CSL MRS plus the combined MRS, and the CSL MRS plus the ComBat-preprocessed MRS in the held-out FHS-UM dataset. Likelihood ratio tests were then used to compare each of the two-MRS models to that CSL-only model, with the resulting p-values indicating whether either of these alternative scores provided additional predictive benefit. MRS evaluation in the REGICOR case-control used logistic regression models, adjusting for the same sets of covariates where possible, though traditional biochemical risk factors were only available in discrete low vs. high categories.

The biology underlying the CSL model was evaluated through a series of enrichment tests using the component CpG loci and annotated genes. Gene ontology-based enrichment analysis of each cohort-specific model was performed using the gometh function from the *missMethyl* package for R^25^. This procedure uses gene annotations for CpGs from the HumanMethylation450 microarray annotation from Illumina (v1.0 B2). Enrichment analysis is then performed for each gene ontology category using Wallenius’ noncentral hypergeometric distribution to account for inconsistent representation of CpG sites across genes. The overall merged set of CpGs included in the final CSL model was then tested for enrichment in transcription factor binding sites using HOMER tool^26^. CpG loci (with respect to genome build hg19) were provided as inputs, with 200 base-pair windows and repeat-masked sequences.

### Genomic risk score calculation

Imputed genotype data for WHI were retrieved from dbGaP (accession: phs000746.v2.p3. Variants were filtered for imputation R-squared > 0.3, and annotated with rsIDs, loci, and allelic information using the 1000 Genomes Phase 3 download from dbSNP (download date: April 13, 2018). Weights for the genetic risk score calculation (6,630,151 variants) were based on the genome-wide CVD score developed by Khera et al^15^. We note that these scores were developed only for populations of European descent, and thus are not optimized for the mixed-ancestry WHI population. GRS were then calculated as the weighted sum of allelic dosages, normalized by the number of relevant SNPs available. Genotype data processing and GRS calculation were performed using PLINK 2.0.

### Risk score interaction analysis

Interaction analysis was performed using similar Cox regression models to those above, adjusting for the “basic” set of covariates and using robust standard error estimates. To facilitate visual comparisons, main-effect regressions for the MRS were fitted within risk strata defined by the FRS or GRS separately in each dataset. To obtain overall interaction effect estimates, an interaction between MRS and either FRS or GRS was introduced into a combined regression including all datasets, while allowing stratified baseline hazards (strata() argument to the coxph function). We note that main effects in the interaction analysis are biased upwards since the regression datasets were used for training the MRS. Regressions assessing the GRS excluded non-white participants to match the ancestry used to develop the CVD score^15^.

For quasi-replication of these associations in the REGICOR dataset, stratified logistic regressions were used to discriminate MI cases from controls using the MRS, while adjusting for estimated cell count fractions as well as two SVA components (as in the main REGICOR models). In the absence of continuous values for blood pressure and lipids, an empirical risk function was generated by first performing a logistic regression on the following cardiovascular risk factors: age, sex, estimated cell count fractions, BMI, diabetes, smoking status, hyperlipidemia (binary), and hypertension (binary), along with two SVA components. Predicted risks based on this model were then used to stratify subjects into four risk groups by evenly splitting the range of predicted risks into four segments (thus resulting in strata based on raw risk, rather than percentiles).

## Results

### Cross-study learner model development

Epigenomic model development was performed in three cohorts, including the Women’s Health Initiative (WHI), Framingham Heart Study Offspring Cohort (FHS), and Lothian Birth Cohort 1936 (LBC). The FHS dataset was divided into two functionally separate groups (FHS-JHU and FHS-UM) based on differences in subject selection and geographic location of laboratory methylation analysis (see Methods). Further population details can be found in Table 1.

**Table 1:** Baseline parameters of the populations used for model development

| Study/Subset | WHI | FHS-JHU | LBC | FHS-UM |
| --- | --- | --- | --- | --- |
| Sample size | 2023 | 484 | 818 | 2103 |
| Age | 65 (59-70) | 71 (64-77) | 69 (68-70) | 64 (59-71) |
| Sex (female) | 2023 (100%) | 145 (30%) | 406 (50%) | 1270 (60%) |
| Ancestry |  |  |  |  |
| % European | 959 (47%) | 484 (100%) | 818 (100%) | 2103 (100%) |
| % African American | 651 (32%) | 0 (0%) | 0 (0%) | 0 (0%) |
| % Hispanic | 413 (20%) | 0 (0%) | 0 (0%) | 0 (0%) |
| Body mass index | 29.1 (25.5-33.3) | 28.2 (25.5-31.3) | 27.5 (24.9-30.3) | 27.4 (24.3-31) |
| LDL cholesterol | 150 (126-175) | 88 (73-107) | 118 (89.5-150.3) | 107 (87-128) |
| HDL cholesterol | 51 (43-60) | 49 (40-60) | 56.1 (47.2-68.3) | 56 (45.8-69) |
| Triglycerides | 127 (92-177) | 101.5 (75-141.2) | 128.4 (97.4-171.2) | 102 (73-142) |
| Fasting glucose | 96 (88.6-108) | 106 (97-116) | Unavailable | 100 (94-109) |
| Systolic blood pressure | 131 (120-143) | 130 (117-143) | 148.7 (137-161.3) | 126 (116-138) |
| # CVD events |  |  |  |  |
| Prior only | 0 | 127 | 70 | 112 |
| Incident only | 1009 | 67 | 133 | 146 |
| Prior and incident | 0 | 58 | 164 | 34 |

\* Continuous values shown as: median (interquartile range)
WHI = Women’s Health Initiative, FHS-JHU = Framingham Heart Study Offspring Cohort (Johns Hopkins University subset), LBC = Lothian Birth Cohorts 1936, FHS-UM = Framingham Heart Study Offspring Cohort (University of Minnesota subset)

Fig. 1 outlines the computational workflow. Briefly, a cross-study learning (CSL) model was developed by training time-to-event elastic net regressions on three of the datasets, while holding out the FHS-UM subset for evaluation. Next, a model trained on all four datasets was subject to external replication in the REGICOR study. CSL model CpGs were characterized as to their potential biological function, and model performance was assessed across strata of alternative cardiovascular risk metrics.

**Figure 1:**
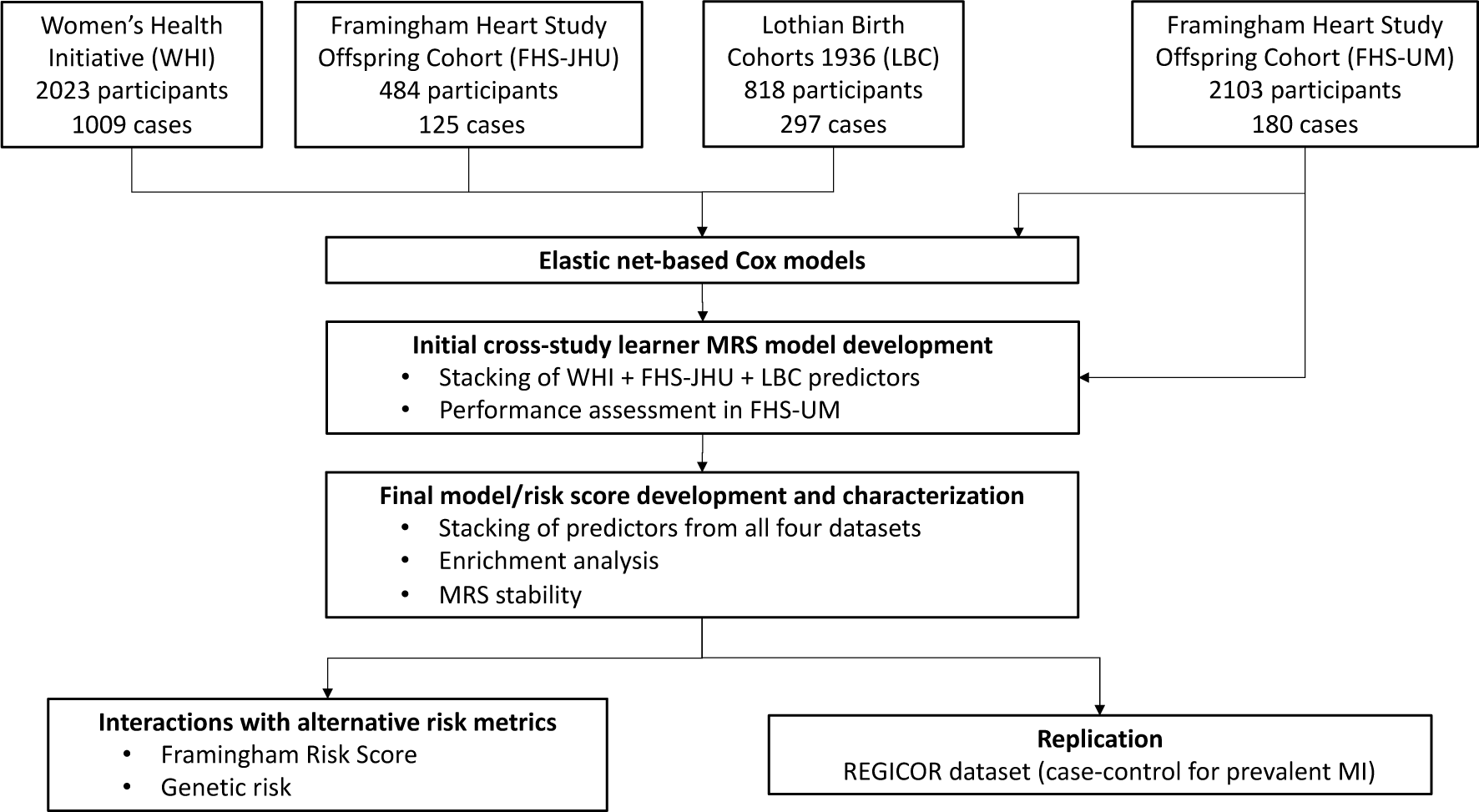
Computational workflow for MRS development and evaluation. The initial MRS was trained in three cohorts with FHS-UM held out to evaluate performance. The final MRS was then trained using all four datasets and examined for biological significance, before testing for prevalent MI discrimination in an independent cohort and assessment of interactions with genetic and traditional risk scores.

**Figure 2:**
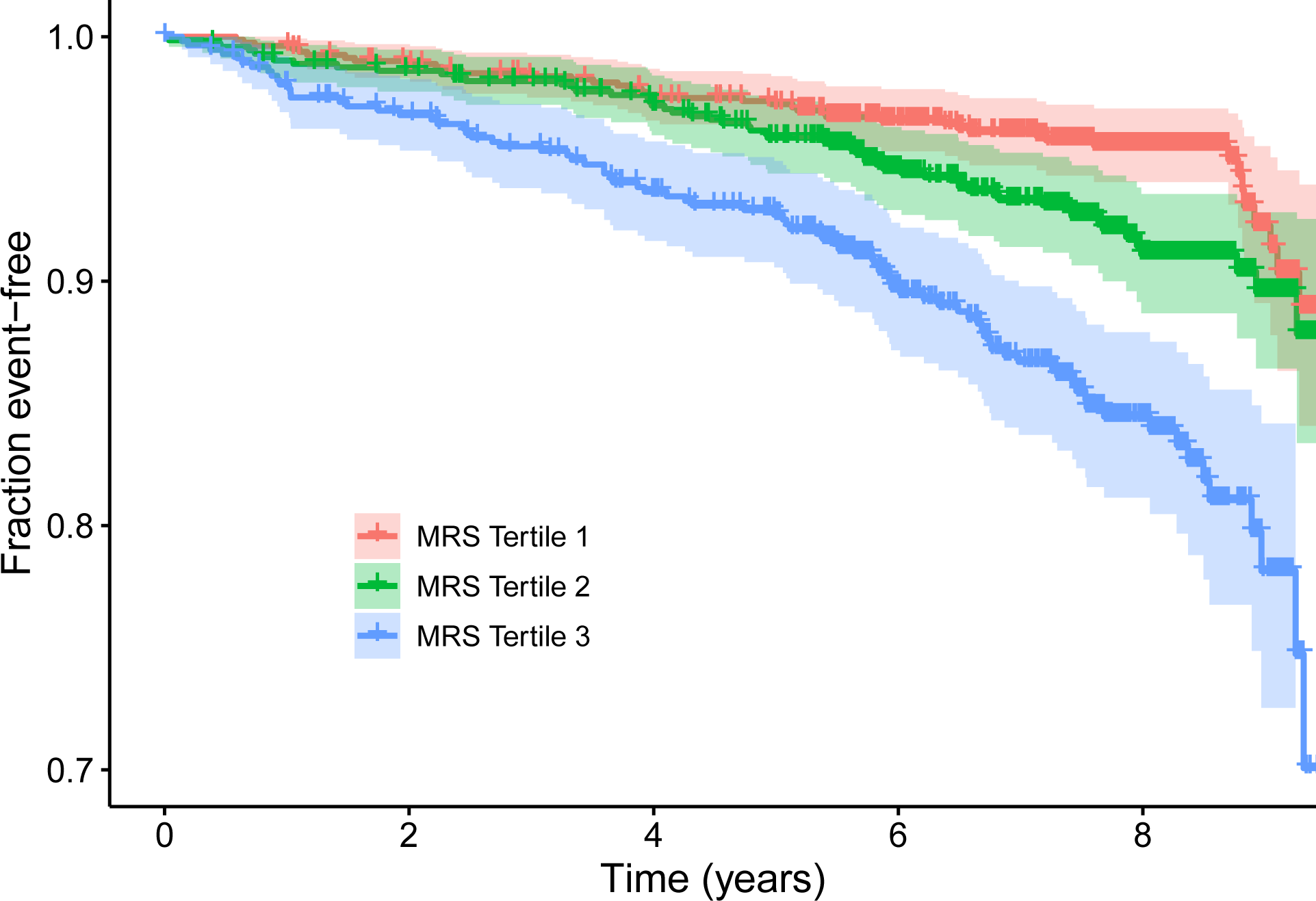
Kaplan-Meier survival curves in the held-out FHS-UM dataset. Individual curves correspond to tertiles of the initial (3-dataset) MRS. Vertical ticks correspond to censored observations, and colored bands represent 95% confidence intervals for tertile-specific survival curves. X-axis is limited to the time span in which at least 50 uncensored observations remained for each tertile (3275 days).

The initial predictor was developed by training individual penalized Cox proportional hazards regression models (single-study learners, or SSLs) in each of the three training cohorts (WHI, FHS-JHU, and LBC). Predictions from these models were aggregated through a “stacking” method, in which the outcomes and model predictions from each of the individual datasets are combined, and a regression is used to assign weights to each of the model predictions (see Methods). This procedure led to FHS-JHU dropping out of the ensemble model, with weights for this initial predictor as follows: 0.57 (WHI), 0.0 (FHS-JHU), and 0.43 (LBC). This result means that FHS-JHU did not generalize (i.e. to WHI and LBC) as well as the other two components models.

### Assessment in held-out FHS subset

Stacking of the three initial predictors resulted in model weights of 0.57, 0, and 0.43 for WHI, FHS-JHU, and LBC, respectively (i.e. the FHS-JHU sub-model did not ultimately contribute to the initial ensemble model). The resulting ensemble predictor was evaluated using robust Cox proportional hazards models in FHS-UM, showing strong associations with incident CVD in an unadjusted model (HR=1.58, 95% CI: 1.37-1.83), which was attenuated partially through adjustment for standard covariates (age, sex, and estimated cell type fractions; HR=1.28, 95% CI: 1.10-1.50) as well as CVD risk factors (HR=1.29, 95% CI: 1.09-1.51). Results for the unadjusted model and three sensitivity models are shown in Table 2. These results were robust to sensitivity analyses excluding all individuals who experienced prior CVD events (Supp. Table S1), and produced similar results when incident event status was analyzed as a binary outcome using logistic regression (p = 3.3e-4).

**Table 2:** MRS performance in held-out FHS subset

| Model | HR per s.d. MRS<sup>a</sup> | P-value |
| --- | --- | --- |
| Unadjusted<sup>1</sup> | 1.58 | 5.4e-10 |
| Basic<sup>2</sup> | 1.28 | 2.0e-03 |
| Plus risk factors<sup>3</sup> | 1.29 | 2.7e-03 |
| FRS only<sup>4</sup> | 1.36 | 2.0e-05 |

<sup>a</sup> Estimated hazard ratio per standard deviation of the methylation-based risk score
<sup>1</sup> No covariates
<sup>2</sup> Adjusted for age, sex, and estimated cell type fractions
<sup>3</sup> Additionally adjusted for BMI, LDL, HDL, SBP, diabetes status, and current smoking
<sup>4</sup> Adjusted for Framingham Risk Score only

Results from comparison of CSL performance to models trained on combined datasets (either naive combination or including preprocessing using ComBat) are shown in Supp. Fig. S1. The ComBat-preprocessed model had modestly higher hazard ratios in FHS-UM, while relative differences with the combined model depended on the covariates included. However, likelihood ratio tests using the Basic model covariates (age, sex, and cell type fraction-adjusted) did not reveal a strong added predictive benefit of either the combined (p = 0.58) or ComBat (p = 0.08) risk scores over that using only the CSL.

### Final CSL model characterization

The stacking regression in the final CSL model gave the most weight to WHI (0.48) and LBC (0.38), while retaining nonzero weights for FHS-JHU (0.06) and FHS-UM (0.08). This result indicates that the WHI and LBC-trained models were better able to generalize to the other cohorts than vice versa. There was very little overlap of specific CpG sites across cohort-specific models, with a maximum of 13 CpGs shared between two models (WHI and FHS-UM) and no CpGs shared between three or more models (Fig. 3a). Despite this lack of site-specific overlap, there was broad agreement for three of the four component SSL models at the level of enriched biological processes, with all except FHS-JHU enriched most strongly for proximity to genes involve in homophilic cell adhesion (Fig. 3b). MRS component CpGs tended to be found in similar genomic loci to the overall set of variable CpGs, and were enriched in gene bodies and depleted in CpG islands compared to the full microarray CpG set. However, MRS CpGs did show a modest enrichment in and around CpG islands compared to the set of variable CpGs (Fig. 3d). To seek more clarity as to potential biological mechanisms represented by the MRS, the HOMER tool was used to calculate enrichment of transcription factor (TF) binding motifs in the MRS component CpG sites. Using the union of all individual SSL CpG sites as input, no strong enrichments were found (all q-values >0.5).

**Figure 3:**
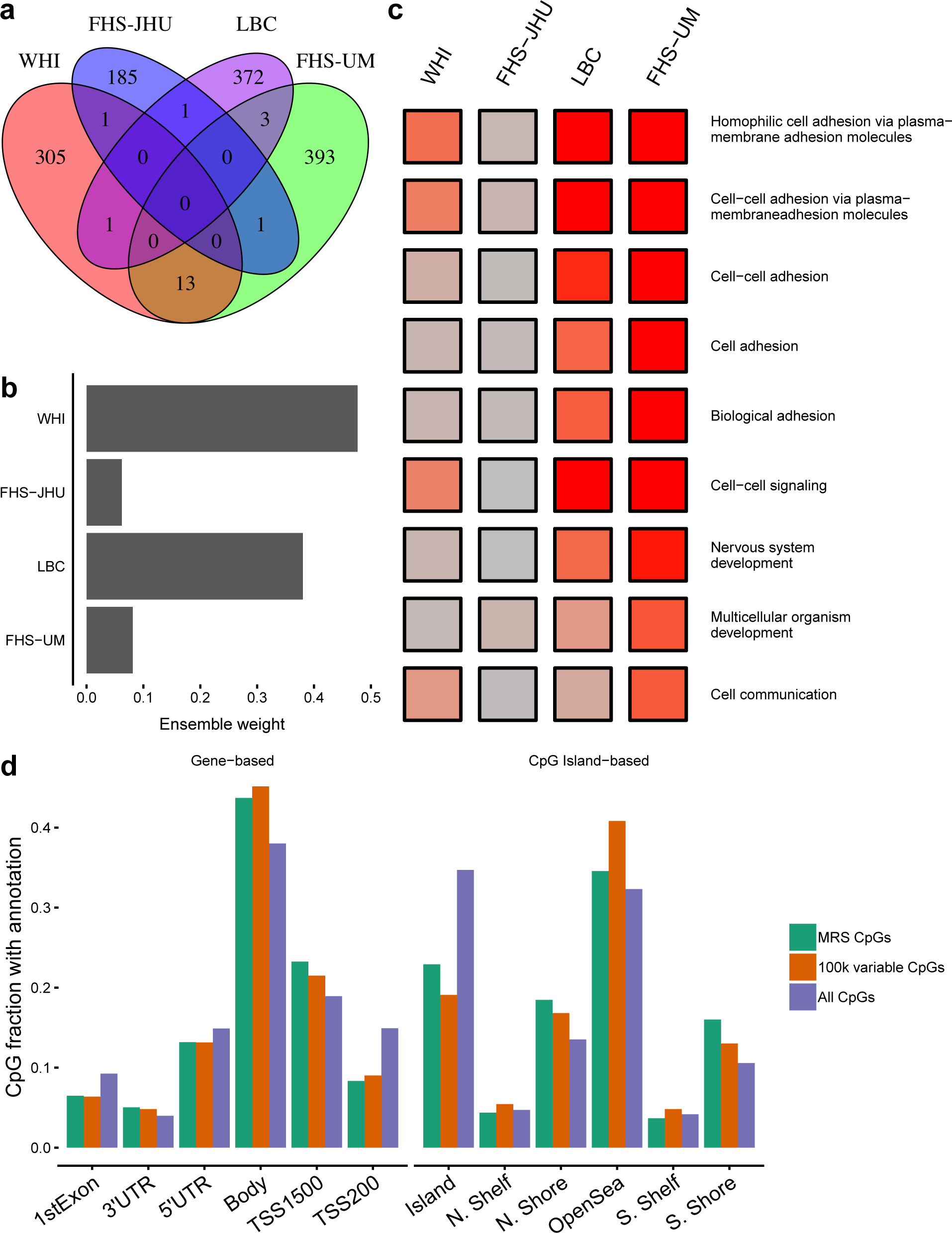
Characterization of the final CSL model. a) Overlap of CpG sites in the four individual predictors constituting the final model. b) Study-specific weights for constructing the ensemble model (derived from the “stacking” regression). c) Results from Gene Ontology-based enrichment analysis using genes annotated to SSL component CpGs. All GO terms with false discovery rate < 0.001 in any cohort are shown, and colored according to -log(p-value) for enrichment in each SSL. Values were cut at -log(p) = 20 for visualization purposes. d) Proportion of CpGs in the full set of CSL CpGs (union of CpG sets in each component SSL) compared to the 100,000 most variable CpGs (as used in SSL model development) and the full set of available CpGs. Groupings according to both gene-based and CpG island-based CpG annotations are shown.

To better understand the stability of the risk score over time, intraclass correlation coefficients (ICCs) were calculated for two sets of grouped samples: 26 technical replicates from FHS and approximately 1000 longitudinal samples (across 3 visits, or about 6 years total) from LBC (Supp. Table S2). The technical replicates showed an ICC of 0.85, while the longitudinal samples showed an ICC of 0.68. As would be expected, the ICC for samples closer in time (Waves 1 & 2; ICC = 0.69) were higher than that for samples more distant in time (Waves 1 & 3; ICC = 0.61). Based on the observation of imperfect stability of the MRS over time as well as the partial attenuation in predictive power after adjustment for age, its component CpGs (the 1305-element union of all CpGs in any of the four individual SSL models) were examined for overlap with established epigenetic age metrics. While no enrichment was seen for the original cross-tissue DNAm age from Horvath^27^, strong enrichment was seen for the morbidity-directed PhenoAge^5^ (9 of 513 CpGs; p=2.3e-5) and especially the blood-specific aging marker from Hannum et al.^28^ (13 of 71 CpGs; p=5.9e-21). We note that these overlaps do not constitute a major fraction of either CpG set, but are nonetheless highly statistically significant. The PhenoAge metric is based on some known cardiovascular risk factors (e.g. C-reactive protein) and is known to associate with CVD, but is not trained in any of the cohorts included here.

### Discrimination in myocardial infarction case-control

As one form of replication, the MRS was investigated for its discriminative performance in a nested case-control for prior myocardial infarction in the REGICOR cohort (Table 3; cohort description in Supp. Table S3). Though this dataset contained prevalent (rather than incident) events, its matching of sex and age allowed an evaluation free of potential confounding by these factors. The MRS was able to discriminate cases and controls in both unadjusted (odds ratio = 1.79, p = 6.33e-6) and, to a lesser degree, risk factor-adjusted models (odds ratio = 1.61, p = 0.019). Given the relatively large confidence intervals, there was no meaningful difference in odds ratios across modeling strategies (Combined, ComBat, and CSL) for any of the adjustment models.

**Table 3:** Results from replication in REGICOR MI case-control

| Model | ComBat | Combined | CSL |
| --- | --- | --- | --- |
| Unadjusted | 1.79 [1.39-2.31] | 1.86 [1.45-2.38] | 1.83 [1.41-2.37] |
| Basic | 2.16 [1.58-2.93] | 2.12 [1.57-2.87] | 2.14 [1.58-2.89] |
| Plus risk factors | 1.76 [1.22-2.54] | 1.66 [1.15-2.4] | 1.61 [1.11-2.34] |

\* Results are presented as: OR per s.d. MRS [95% CI]
\* Model covariates as in Table 2
\* All models above are adjusted for two surrogate variable analysis (SVA) components.

### Interactions with alternate risk metrics

To understand how the present risk score interacts with other established CVD risk metrics, the performance of the MRS was re-evaluated after stratifying individuals by risk scores reflecting either demographic and biochemical features (Framingham Risk Score), or genetic variants (based on Khera et al. 2018). First, the marginal effects of these risk scores were confirmed in each population. The Framingham Risk Score (FRS) was strongly predictive in WHI and FHS, while surprisingly showing no association with CVD incidence in LBC (Supp. Table S4). The genetic score was evaluated in WHI, demonstrating a moderate association with CVD (odds ratio per standard deviation = 1.28, p = 1.1e-6).

In Cox models using baseline hazards stratified by study and performed using the final four-study MRS, it appeared that the MRS was more effective in those in lower “traditional” risk strata (according to the FRS; Fig. 4). As a sensitivity analysis, the cohorts were fully stratified into separate models, in which this pattern was visually clear in WHI and FHS-JHU (Supp. Fig. S2). The pattern did not appear in LBC, although we note that the Framingham Risk Score also did not show a “main effect” for predicting incident CVD in this cohort. A similar pattern appeared with respect to genetic risk in WHI (European ancestry participants only based on the formulation of the relevant risk score), in which maximum MRS performance was achieved in the lowest alternative risk stratum. Supplementing these visual comparisons, combined Cox regressions across all cohorts (allowing for different baseline hazards across studies) showed a strong MRS-FRS interaction effect (7% reduction in HR for the MRS per 10% increase in FRS; p = 8.27e-05), while that for the MRS-GRS interaction did not reach nominal statistical significance (2% reduction in HR for the MRS per standard deviation increase in GRS; p = 0.719).

**Figure 4:**
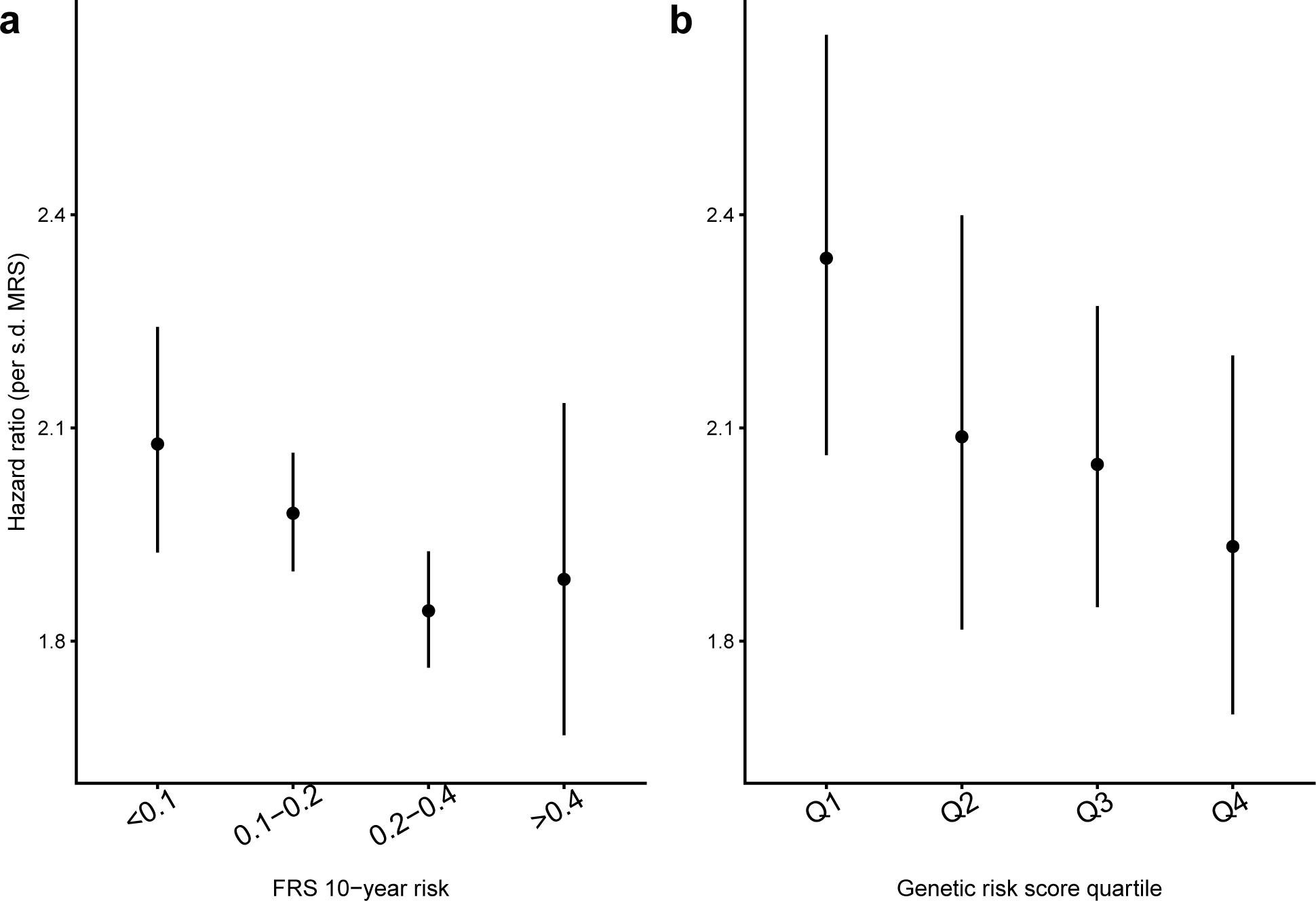
Interactions of MRS with other biomarkers of CVD risk. a) Hazard ratios for the MRS within subsets of 10-year generalized CVD risk according to the Framingham Risk Score. b) Hazard ratios for the MRS within quartiles of a genetic cardiovascular risk score (in white WHI participants only). Hazard ratios are estimated using the final MRS, which was trained using each of these datasets. Cox regressions included stratum-specific baseline hazards and were adjusted for age, sex, and estimated cell subtype fractions. Error bars represent standard errors for the hazard ratio estimates.

To explore the clinical potential of these interactions further, we returned to the initial MRS (trained in 3 datasets with FHS-UM held out). The FHS-UM dataset was filtered to include only participants with lower CVD risk based on the FRS (<10% estimated 10-year risk). Within this lower-risk subset, participants in the upper MRS quintile had more than double the risk of the remainder of the participants: 7% (12/176) versus 3% (19/701).

FRS could not be calculated in the REGICOR dataset, as not all risk factors were available as continuous values. However, stratified models replicated the observation of greater MRS discrimination in the lowest alternative risk stratum. An empirical risk function was generated through logistic regression of MI status on cardiovascular risk factors (age, sex, BMI, diabetes, smoking status, hyperlipidemia (binary), and hypertension). Predicted MI risk using this model was used to stratify subjects into four risk groups, with MRS odds ratios (per standard deviation) of 4.49 in the lowest-risk group versus 1.20 in the highest-risk group. More detailed results from these analyses are shown in Supp. Table S5.

## Discussion

Epigenetic signatures of cardiometabolic diseases and aging in general are being actively explored as biomarkers of disease risk that are potentially modifiable and reveal underlying biological mechanisms. Here, in a novel application of a cross-study ensembling method, we introduce a DNA methylation-based score specific to cardiovascular disease risk. The model performs similarly to one trained on a direct combination of the component datasets, and may be most strongly predictive in individuals predicted to be at lower risk based on traditional risk factors.

We opted to use cross-study learning to train our risk model based on the expectation that differences across cohorts (e.g. demographic, behavioral) may contribute to heterogeneity in both the marginal distribution of the CpG features and the conditional distribution of the CVD outcome. Under these conditions, the generalizability of a single-study predictor is often obscured or overstated^29,30^. The performance of the CSL model was similar to that of models trained on the merged cohorts with or without batch adjustment via ComBat. This suggests that the assumptions made by these direct combination strategies (i.e. that the heterogeneity structure can be captured by variation in the marginal effects of each CpG site) are met. In practice, this underlying structure is unknown, and we highlight that the CSL was able to produce similar gains in predictive accuracy without making specific assumptions.

In assessing the stability of the MRS, we observed reasonable reproducibility between technical replicates (ICC=0.85). ICCs for LBC subjects over time were somewhat lower (ICC=0.68), which is to be expected due to not only changes in environment, but also the known epigenetic evolution with age that we observed to be enriched in the components of our score. Furthermore, this value is at the upper end of the range of single-CpG repeatability measurements over time calculated in the combined Lothian Birth Cohorts (1921 and 1936)^31^. These ICC values suggest an imperfect but usable reproducibility of the MRS, and an aggregate marker that is fairly robust considering the low replicability that has been observed for individual sites in technical replicates (general median ICC of 0.3 and mode of 0.75 in a “high reliability” cluster)^32^.

The enrichment of the MRS component CpGs for proximity to genes related to cell-cell adhesion (in all subsets except FHS-JHU) is indicative of the underlying biological mechanisms. As we have previously observed in the WHI and FHS cohorts, it appears that immune activation is central to the prognostic information contained in leukocyte DNA methylation^14^. For example, epigenetic processes have been shown to be involved in the activation and increased adhesion of monocytes in response to environmental insults and metabolic stress, though these have been explored primarily in relation to histone modifications^33^. Our results provide preliminary support for an attractive model in which a methylation-based score could act as a monitor of cumulative stress in leukocytes and their corresponding activation towards a more atherogenic state.

Existing epigenetic scores have shown varying strength in predicting incident cardiovascular disease. An early investigation examined blood-based methylation in LINE-1 elements, finding strong associations of global hypomethylation with prevalent and incident ischemic heart disease global (LINE-1), though additional reports showed opposite associations of methylation at repetitive elements with CVD^34^. Guarrera et al. developed a biomarker for MI based on global LINE-1 and ZBTB12 gene methylation that provided a modest net reclassification index improvement (0.23-0.47) compared to traditional risk factors only. Multiple epigenetic aging metrics, though not developed specifically for CVD, have been shown to predict incident CHD, including PhenoAge (odds ratios from 1.02 to 1.08) and GrimAge (hazard ratio = 1.07, adjusted for age and technical factors)^5,24^. While these associations are statistically significant, they do not represent clinically meaningful improvements in discrimination. Our observed hazard ratio of 1.28 (Basic model in the held-out FHS-UM dataset) indicates that this MRS may be closer to clinical relevance. We note that our component CpG sites overlap strongly with those of these established epigenetic metrics including PhenoAge, suggesting that it captures some of the same biological patterns. However, the mechanistic significance of the specific methylation signals captured by these aging-related metrics, whether as markers of epigenetic regulation breakdown or the work of an “epigenetic maintenance system”, is still unclear^27,35^.

In examining the potential clinical utility of an novel risk score for CVD, it is important to understand to what extent it is redundant with or complementary to existing risk metrics. This type of cross-metric analysis can be clinically relevant, as demonstrated for example in a recent investigation exploring the interaction between genetic and lifestyle-based risk prediction for dementia^36^. Here, we saw a pattern of improved epigenetic risk prediction in individuals whose cardiovascular risk based on traditional metrics (here, the Framingham Risk Score) was low. This pattern replicated in the REGICOR dataset (though FRS could not be directly calculated), with improved MRS discrimination in lower-risk subjects based on an empirical risk function. While these associations are preliminary, they suggest that an epigenetic risk score could help identify higher-risk individuals who otherwise would not have been detected by other metrics. While we did not identify any robust patterns of differential MRS performance in strata based on a genetic cardiovascular risk score, there may have been lower power to detect any such patterns from the outset given the modest discriminatory performance of the GRS in WHI.

Multiple limitations should be acknowledged. While lymphocytes are known to be important in CVD pathogenesis, there is likely additional biological signal in other CVD-relevant tissues not examined here. Additionally, the present definition of CVD was chosen to balance specificity of CVD subtypes with sample size, but this balance could be altered to focus on more specific disease subtypes (e.g. myocardial infarction) or a broader definition of CVD (e.g. including heart failure). Finally, while the REGICOR dataset provided an important age- and sex-matched case-control setting for replication of the MRS, this work would benefit from future replication in an independent cohort enabling assessment of incident disease.

In sum, we have developed an epigenetic risk score for cardiovascular disease that provides additional predictive power beyond existing risk measures, and may show improved performance in populations otherwise designated as low-risk. Furthermore, we have shown a novel application of a cross-cohort ensembling method that may provide significant value to future investigations in genomic epidemiology.

## Data Availability

See Methods and Supplementary Methods for locations and accession numbers of controlled-access data (dbGaP and EGA).

## Acknowledgements

We thank all LBC1936 study participants and research team members. The LBC1936 is supported by Age UK (Disconnected Mind program) and the Medical Research Council (MR/M01311/1). Methylation typing in LBC1936 was supported by Centre for Cognitive Ageing and Cognitive Epidemiology (Pilot Fund award), Age UK, The Wellcome Trust Institutional Strategic Support Fund, The University of Edinburgh, and The University of Queensland. LBC1936 work was conducted in the Centre for Cognitive Ageing and Cognitive Epidemiology, which is supported by the Medical Research Council and Biotechnology and Biological Sciences Research Council (MR/K026992/1), and which supported IJD.

